# How many medications do doctors in primary care use? An observational study of the DU90% indicator in primary care in England

**DOI:** 10.1101/2020.06.16.20132654

**Authors:** Chiamaka Chiedozie, Mark Murphy, Tom Fahey, Frank Moriarty

**Affiliations:** HRB Centre for Primary Care Research, Department of General Practice, Royal College of Surgeons in Ireland; School of Pharmacy and Biomolecular Sciences, Royal College of Surgeons in Ireland

**Author notes:** **Corresponding author:** Frank Moriarty, School of Pharmacy and Bimolecular Sciences, Royal College of Surgeons in Ireland, Ardilaun House Block B, Dublin 2, Ireland. **Principal investigator statement:** This study did not have a Principal Investigator, as it reports an analysis of publicly available administrative data.

**Keywords:** drug utilisation, primary care, general practice, prescribing costs, quality of care

## Abstract

**Aim:** To apply the DU90% indicator (the number of unique drugs which make up 90% of a doctor’s prescribing) to GP practices prescribing in England to examine time trends, practice-level variation, and relationships with practice characteristics

**Method:** This is an observational cohort study of all general practices in England. It utilises publicly available prescribing data from the National Health Service (NHS) Digital platform for 2013-2017. The DU90% was calculated on an annual basis for each practice based on medication BNF codes. Descriptive statistics were generated per year on time trends and practice-level variation in the DU90%. Multi-level linear regression was used to examine the practice characteristics (relating to staff, patients, and deprivation of the practice area).

**Results:** A total of 7,623 GP practices were included. The mean DU90% ranged from 130.1 to 133.4 across study years, and variation between practices was low (with a 1.4 fold difference between the lowest and highest 5% of practices). A range of medications were included in the DU90% of virtually all practices, including atorvastatin, levothyroxine, omeprazole, ramipril, amlodipine, simvastatin and aspirin. A higher volume of prescribing was associated with a lower DU90%, while having more patients, higher proportions of patients who are female or aged 65+, higher number of GPs working in the practice, and being in a more deprived area were associated with a higher DU90%.

**Conclusion:** GP practices typically use 130 different medications in the bulk of their prescribing. Increasing use of personal formularies may enhance prescribing quality and reduce costs.

1. *What is already known about this subject:*
  - Prescribers can develop expertise in a finite number of drugs, hence use of a personal formulary is recommended and may enhance prescribing quality
  - The drug utilisation 90% (DU90%), a measure of the number of drugs that comprises 90% of prescribing volume, has not been applied to prescribing in England to date.
2. *What this study adds:*
  - On average, 130 medicines make up 90% of prescribing in GP practices in England.
  - Practices with a lower DU90% tended to have lower prescribing costs and less prescribing considered low-value care.
  - Rationalising prescribing to focus on a smaller group of medicines may enhance quality and reduce costs.

## Introduction

Prescribing is the most common healthcare intervention in developed countries, however decisions to prescribe or continue a medication for a patient are complex, with many factors that must be considered. These include a medication’s indication, dosage, frequency, duration of use, adverse effects, contraindications and interactions with other medications or conditions an individual may have.[1] Doctors can develop expertise in only a certain number of drugs, and so may have to consult information sources when considering others they are less familiar with. For this reason, the World Health Organisation’s Guide to Good Prescribing suggests that prescribers should develop a personal list (i.e. drugs they have chosen to prescribe regularly which they are familiar with that are their priority option for given indications).[2] This should support the selection of the right drug for each patient in the relatively short time that may be available to consider this during a consultation. While a small number of preferred drugs may be feasible for specialist prescribers, this may prove challenging for general practitioners (GPs) as generalists prescribing a wide range of treatments,[3] and who often manage all medications for patients, including those initiated by specialists in secondary care.[4–6]

Use of a relatively limited number of core drugs, which a prescriber is an expert in, may reduce task complexity in treatment decisions and potentially be associated with a higher quality prescribing and reduced errors.[7, 8] The availability of many alternatives with similar therapeutic effects can compromise patient safety through potential confusion between drugs within a class but with different properties or dosage.[9] Mitigating this risk of confusion is one basis for developing a core drug list for undergraduate and postgraduate prescribing education in the UK.[10, 11] Some countries have implemented restricted formularies or essential medicines lists on this basis, such as the ‘Wise List’ in Sweden and local formularies in the UK,[9, 12, 13] to make it facilitate clinicians in prescribing the most effective, safe and appropriate medications for their patients.[14]

The drug utilisation 90% (DU90%) is a measure recommended by the World Health Organisation (WHO) for drug utilisation research, and represent the number of unique drugs which make up 90% of a doctor’s prescribing.[3] The number, as well as the specific drugs, in this segment, may serve as simple indicators of the quality of drug prescribing. It has been used to provide feedback to GP practices on their prescribing patterns for quality assurance.[3, 12] A DU90% segment with a large number of drugs may indicate use of medications which a prescriber sees infrequently, which may increase the risk of sub-optimal prescribing. Similarly, a segment with limited number of drugs may indicate more rational prescribing.[15] Wettermark et al. assessed the usefulness of DU90% as a tool for indicating prescribing quality, and found high acceptance of this among GPs in Sweden who regarded DU90% profile as clear, relevant and useful for improving prescribing quality.[16]

The aims of this study are:

- To characterise the DU90% indicator in prescribing data from GP practices in England, by assessing time trends, variation between practices, and relationship with practice characteristics.
- To assess the relationship between DU90% and prescribing deemed to be low-value care and prescribing costs.

## Methods

### Study Design, Setting and Participants

This is a repeated cross-sectional study of GP practices in England and is reported in line with the Strengthening the Reporting of Observational Studies in Epidemiology (STROBE) statement.[17] It used publicly available prescribing data available from the National Health Service (NHS) Digital platform. This provides monthly statistics of prescribing of different medicines aggregated at the level of GP practices for all practices in England. The data relate to NHS prescriptions issued by any prescribing staff in GP practices in England and dispensed in any community pharmacy in the UK, and private prescriptions are not included. The study period was over five years from January 2013 to December 2017. Atypical practices were excluded, i.e. those with <1,000 registered patients, without a Quality and Outcome Framework (QOF) Score, or contributing data in fewer than three consecutive years (either no data or on less than 1,000 prescriptions in a year) to allow assessment of time trends. As this study used publicly available data aggregated at the GP practice-level, ethical approval was not required.

### Variables

The DU90% indicator was calculated on an annual basis for each GP practice. The number of prescriptions in each year per medicine was summarised at the practice level. Unique medicines were identified using the first nine digits of the British National Formulary (BNF) code, which identifies each chemical substance. In line with previous studies which excluded products without a WHO Defined Daily Dose,[3, 15, 18] certain non-oral pharmaceutical products were excluded before analysis based on their BNF codes (see eTable 1). Medicines were arranged in descending order of total number of prescriptions for each medicine within a practice and the DU90% calculated represents the number of medicines where the cumulative sum of prescriptions is >90% of all prescriptions issued by the practice.

Characteristics of GP practices were of interest as explanatory variables potentially associated with the DU90%. These included what Clinical Commissioning Group (CCG) the practice is part of, the deprivation of the area a practice is located in, number of registered patients and age and sex distribution of registered patients, quality of care indicated by the QOF score, and practice workforce (i.e. the number of full-time equivalent GPs, age and sex distribution of GPs, and whether the practice has a registrar GPs).[19]

### Data sources

Prescribing data from NHS England captures the number of prescription items for each product that was dispensed in the specified month per practice. It also includes the product’s BNF code, the quantity of dosage units dispensed per product, the net ingredient cost (NIC), and the actual cost per product. GP practice workforce, registered patient data, and overall QOF performance for each year for included practices were downloaded from the NHS Digital website and summarised at the practice level. For deprivation, the index of multiple deprivation (IMD) for 2015 is provided for geographic areas (lower-level super output areas or LSOA) by the Department for Communities and Local Government. The index captures the following dimensions of deprivation: income, employment, education, health, crime, access to housing and services, and living environment. Practices were assigned the IMD decile of the LSOA they were located in based on their postcode.

### Analysis

Firstly, baseline characteristics of included GP practices were generated. The DU90% per year for individual practices was calculated. To characterise variation in DU90%, we plotted the distribution of practices by their DU90% by year. We also determined the proportion of practices with a DU90% between 2 and 3 SD from the mean, and more than 3 SD from the mean, as well as the extremal quotient per year (winsored at the 5% level). We tested for a linear trend across years using the nptrend function in Stata, and assessed differences between years using a multi-level linear regression model.

We fitted a multi-level linear regression model with random intercepts at the CCG and practice levels. We determined the total variance, as well as the variance between CCGs, between practices, and within practices by calculating the variance partition coefficient. Standardised versions of covariates (i.e. centred on their mean with a standard deviation of 1) were added to this regression model as fixed effects to assess factors associated with the DU90%. Firstly patient and practice characteristics were added, followed by GP workforce characteristics.

Medicines included in the DU90% segment were characterised for both 2013 and 2017. We determined the medicines that feature in the DU90% segment of the most practices by year, and those that constituted the greatest volume of prescriptions within the DU90% by year. We determined the proportion of DU90% medicines across BNF chapters (i.e. physiological systems) and average number of DU90% medicines within common drug classes with multiple drugs (statins, PPIs, ACE inhibitors, ARBs, dihydropyridine calcium channel blockers, beta blockers, SSRIs, penicillins and opioids).

The original DU90% includes an assessment of guideline adherence within the DU90% segment (proportion of medicines in the segment recommended in guidelines). As there is no national guideline for England for recommended medicines, we assessed prescribing of items which the NHS recommends should not be routinely prescribed in primary care due to low effectiveness, poor cost-effectiveness, or being low priority (eTable 2). We summarised both the proportion of all prescribing considered low priority, and the proportion of low priority prescribing within the DU90% for each practice, expressed in terms of prescriptions and costs. Lastly, we assessed whether practices’ DU90% (divided into five groups based on quintiles) was associated with differences in prescribing costs, low priority item prescribing, BNF chapters, and number of agents within common drug classes.

## Results

A total of 7,620 GP practices were included in this study, with the number included during each study year shown in Table 1, along with practice, registered patient, and GP workforce characteristics. Between 153 and 158 medicines made up 90% of overall prescribing for included practices during the study period. At practice level, the mean DU90% ranged from 130.0 to 131.0 across study years, and varied by between 3 fold and 5 fold depending on the year. However, winsoring at the 5% level, the equivalent span was approximately 1.4 across study years (see eTable 3). The vast majority of GP practices had a DU90% within 2 standard deviations of the mean, with 4% falling between 2 and 3 SDs, and less than 1% having a DU90% greater than 3 SDs from the mean. Figure 1 shows the distribution of practice by DU90% across years and also indicates variation was low and remained stable with time.

**Table 1:** Baseline characteristics of included practices.

| Characteristics of practices | n (%)<br>n=7620 |
| --- | --- |
| Patient characteristics |  |
| Total registered patients, median (IQR) | 6366 (3865 to 9715) |
| Percentage of patients aged ≥45 years, mean (SD) | 41.8 (10.2) |
| Percentage of female patients, mean (SD) | 50.0 (2.34) |
| Practice characteristics |  |
| Total annual prescription items, median (IQR) | 92,054 (55,114 to 146,286) |
| Overall QOF score, median (IQR) | 96.3 (92.0 to 98.6) |
| Practices by IMD decile* |  |
| 1 | 1195 (15.7) |
| 2 | 978 (12.8) |
| 3 | 908 (11.9) |
| 4 | 850 (11.2) |
| 5 | 734 (9.6) |
| 6 | 660 (8.7) |
| 7 | 647 (8.5) |
| 8 | 602 (7.9) |
| 9 | 557 (7.3) |
| 10 | 488 (6.4) |
| GP practice workforce** |  |
| GP headcount, mean (SD) | 5.2 (3.5) |
| GP full time equivalents, mean (SD) | 4.5 (3.1) |
| Percentage of GPs aged ≥45 years, mean (SD) | 57.4 (28.4) |
| Percentage of female GPs, mean (SD) | 44.9 (26.0) |
| Practices with a registrar | 1949 (25.6) |
| Practices included per year |  |
| 2013 | 7591 (99.6) |
| 2014 | 7594 (99.7) |
| 2015 | 7596 (99.7) |
| 2016 | 7273 (95.4) |
| 2017 | 7140 (93.7) |

GP, general practitioner; IMD, Index of Multiple Deprivation; IQR, interquartile range; SD, standard deviation; QOF, Quality and Outcomes Framework; \*Missing for 1 practice at baseline; \*\*Missing for 8 practices at baseline

**Figure 1.**
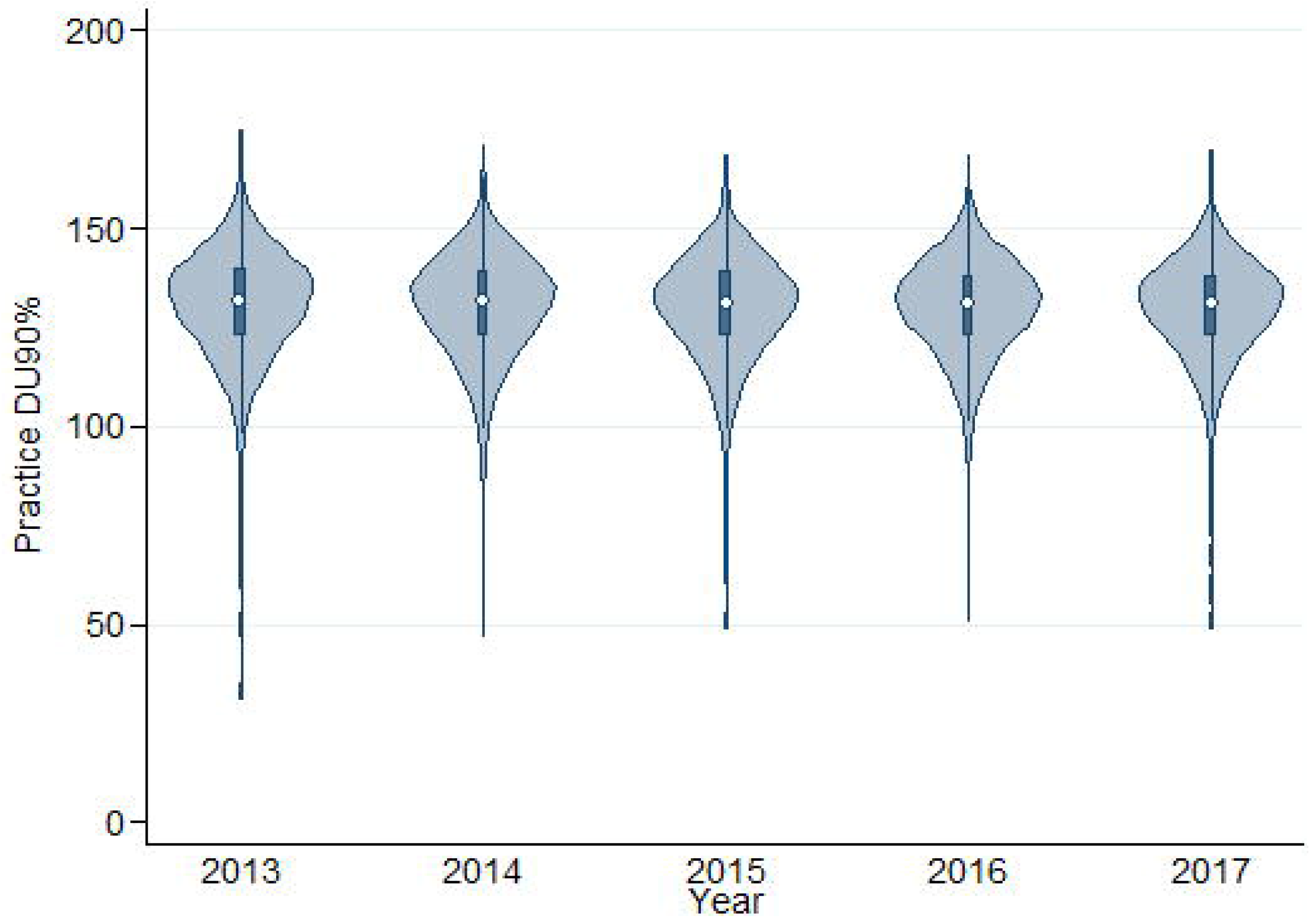
Violin plot showing distribution of GP practices’ DU90% across years.

There was a decreasing linear trend in DU90% across years (p < 0.001). Fitting a mixed linear regression model with random intercepts for CCGs and practices, and study year as a fixed effect (Table 2, Model 1), there was a statistically significant reduction in DU90% across years, although of small magnitude. Based on variance partition coefficients, 24% of variance was between CCGs, 70% between practices, and 6% within practices.

**Table 2:** Characteristics associated with practice DU90% in multi-level linear regression.

| | $\beta$ coefficient (95% confidence interval) | | |
| --- | --- | --- | --- |
|  | Model 1 | Model 2 | Model 3 |
| Year (reference 2013) |  |  |  |
| 2014 | -0.63 (-0.73 to -0.53) | -0.58 (-0.68 to -0.48) | -0.57 (-0.67 to -0.47) |
| 2015 | -0.99 (-1.09 to -0.89) | -1.07 (-1.17 to -0.97) | -1.03 (-1.13 to -0.92) |
| 2016 | -1.10 (-1.20 to -1.00) | -1.26 (-1.37 to -1.16) | -1.20 (-1.32 to -1.09) |
| 2017 | -1.25 (-1.35 to -1.15) | -1.53 (-1.64 to -1.42) | -1.49 (-1.61 to -1.37) |
| Total prescription items* |  | -0.98 (-1.24 to -0.73) | -1.39 (-1.66 to -1.11) |
| IMD decile |  | 0.39 (0.30 to 0.49) | 0.35 (0.26 to 0.44) |
| Overall QOF percentage* |  | -0.07 (-0.13 to -0.01) | -0.07 (-0.13 to -0.01) |
| Registered patients* |  | 3.89 (3.66 to 4.12) | 4.45 (4.19 to 4.70) |
| Proportion of female patients* |  | 0.26 (0.03 to 0.50) | 0.51 (0.27 to 0.75) |
| Proportion of patients aged $\geq 45$ years* | | 1.99 (1.85 to 2.13) | 2.04 (1.89 to 2.18) |
| GP FTEs* |  |  | 0.27 (0.16 to 0.37) |
| GP proportion female* |  |  | 0.03 (-0.04 to 0.09) |
| GP proportion aged $\geq 45$ years* | | | -0.12 (-0.18 to -0.05) |
| Any registrar |  |  | 0.22 (0.07 to 0.37) |
| Variance (VPC) |  |  |  |
| CCG | 38.7 (24.0) | 29.7 (24.7) | 29.4 (24.9) |
| Practice | 112.9 (70.0) | 80.5 (67.2) | 79.1 (67.0) |
| Residual | 9.7 (6.0) | 9.6 (8.0) | 9.5 (8.1) |

\* $\beta$ coefficients represent change in DU90% per change of one SD in covariate value (78,651 in total prescription items, 6.7 percentage points (pp) in overall QOF percentage, 4586 in registered patients, 2.3pp in proportion of female patients, 3 in GP FTEs, 10.3pp in proportion of patients aged 45 years and over, 25.9pp in proportion of female GPs, and 28.7pp in proportion of GPs aged 45 years and over).
GP, general practitioner; IMD, Index of Multiple Deprivation; QOF, Quality and Outcomes Framework; VPC, variance partition coefficient.

Model 2 found higher numbers of registered patients and a higher proportion of patients aged 45 years or over were both associated with higher DU90% (3.89, 95% CI 3.66 to 4.12, more medicines in the DU90% per additional 4,600 registered patients; and 1.99, 95% CI 1.85 to 2.13, higher per 10% increase in proportion of patients aged 45 and over). Higher proportion of female patients and high deprivation were both also associated with increased DU90%, although the magnitude was smaller. Higher total number of prescription items and overall QOF score were both associated with a lower DU90%. When GP workforce characteristics were considered, more GP FTEs or presence of a GP registrar were both associated with higher DU90%, while proportion of GPs aged 45 years and over was associated with a reduction.

Table 3 compares medicines included in the DU90% of practices in 2013 and 2017. The distribution of these medicines across BNF chapters (physiological systems) was similar over the study period, and mean number of agents within common drug classes in a practice’s DU90% was also stable. eTables 4 and 5 list the medicines included in practices’ DU90% segments in 2013 and 2017 respectively, and show that the medicines making up the highest proportions of prescribing were similar in these years. From approximately 1,250 medicines (i.e. chemical substances based on 9 digit BNF codes), 652 different medicines were included in the DU90% across included practices for 2013, and 606 for 2017. In 2017, a range of medicines were included in the DU90% of the vast majority (≥99.0%) of practices across statins (atorvastatin, simvastatin), PPIs (lansoprazole, omeprazole), cardiovascular medicines (ramipril, amlodipine, bisoprolol fumarate, aspirin), inhalers (salbutamol, beclometasone dipropionate), CNS agents (paracetamol, amitriptyline, citalopram) and others (levothyroxine sodium, metformin, folic acid, oral prednisolone). The agents that accounted for the greatest proportion of items prescribed in practices which included them in their DU90% were atorvastatin (4.1%), levothyroxine sodium (3.4%) and omeprazole (3.4%).

**Table 3.** Medication categories within DU90% in 2013 and 2017.

|  | 2013 | 2017 |
| --- | --- | --- |
| **BNF chapters** | **n (%) of prescriptions** |  |
| Gastrointestinal | 93,358 (9.4) | 89,182 (9.6) |
| Cardiovascular | 245,279 (24.7) | 228,782 (24.6) |
| Respiratory | 84,843 (8.5) | 81,301 (8.8) |
| Central nervous system | 262,184 (26.4) | 241,016 (25.9) |
| Infections | 80,851 (8.1) | 67,143 (7.2) |
| Endocrine | 81,299 (8.2) | 83,688 (9.0) |
| Obsgyn, urinary tract disorders | 54,706 (5.5) | 53,129 (5.7) |
| Malignancy and immunosuppression | 4,563 (0.5) | 4,857 (0.5) |
| Nutrition and blood | 25,785 (2.6) | 26,274 (2.8) |
| Musculoskeletal | 58,485 (5.9) | 50,283 (5.4) |
| Anaesthesia | 3,191 (0.3) | 3,169 (0.3) |
| **Drug classes** | **Mean (SD) agents in DU90% per practice** |  |
| Statins | 3.42 (0.65) | 3.50 (0.65) |
| PPIs | 2.74 (0.74) | 2.94 (0.75) |
| ACE inhibitors | 3.60 (0.65) | 3.38 (0.66) |
| ARBs | 3.13 (0.96) | 2.74 (0.73) |
| CCB | 3.02 (0.86) | 2.98 (0.88) |
| BB | 3.65 (0.80) | 3.41 (0.66) |
| SSRIs | 3.96 (0.71) | 3.84 (0.71) |
| Penicillins | 2.90 (0.32) | 2.90 (0.38) |
| Opioids | 4.81 (1.47) | 5.00 (1.40) |

Considering prescribing in 2017, the mean NIC per 1,000 items per practice was £7,835 (SD 1,472), and on average, the DU90% accounted for 67.8% of costs. The low priority items constituted a mean of 0.48% of all prescribing and 0.26% of prescribing in the DU90%, which equated to 2.02% of total costs and 1.19% of DU90% costs. Table 4 divides practices by quintiles of DU90% and illustrates that higher DU90% is related to higher costs, a higher proportion of costs within the DU90%, as well as higher low priority prescribing both overall and within the DU90%, regardless of expression as prescription items or costs. Regarding types of medications, the percentage of cardiovascular, respiratory and endocrine medicines within the DU90% decreased with higher DU90%, while the percentage of CNS and infections medicines increased with higher DU90% (Figure 2). The mean number of agents within each class increased across quintiles. The greatest relative increase was for opioids, PPIs, and calcium channel blockers, whereas the increases were of lesser magnitude for SSRIs, ACE inhibitors and beta blockers. eFigure 1 shows the distribution of practices by the number of agents within drug classes included in their DU90%, split across quintiles of DU90%. eTables 6 and 7 list the individual medicines included in practices’ DU90% segment in 2017 for the bottom and top quintile of DU90%.

**Table 4.** Costs, low priority item prescribing, BNF chapters and drug classes in 2017 across quintiles of DU90%.

|  | 1st | 2nd | 3rd | 4th | 5th |
| --- | --- | --- | --- | --- | --- |
| Net ingredient cost (NIC) per 1,000 prescriptions, £ | 6,274 (1,202) | 6,881 (1,118) | 7,263 (1,157) | 7,909 (1,263) | 8,842 (1,369) |
| DU90% NIC as a percentage of total NIC | 67.4 (4.0) | 67.7 (3.5) | 67.7 (3.2) | 68.2 (2.9) | 68.3 (2.8) |
| Low priority items as a percentage of total items | 0.46 (0.29) | 0.47 (0.28) | 0.46 (0.23) | 0.50 (0.24) | 0.53 (0.21) |
| Low priority items as a percentage of DU90% items | 0.25 (0.28) | 0.25 (0.28) | 0.24 (0.23) | 0.27 (0.24) | 0.28 (0.21) |
| Low priority NIC as a percentage of total NIC | 1.56 (1.20) | 1.93 (1.33) | 2.01 (1.33) | 2.18 (1.27) | 2.46 (1.37) |
| Low priority NIC as a percentage of NIC within DU90% | 0.55 (0.79) | 0.78 (1.12) | 0.84 (1.21) | 0.91 (1.10) | 1.08 (1.23) |
| **Unique agents within classes in DU90%, mean (SD)** |  |  |  |  |  |
| Statins | 3.13 (0.73) | 3.39 (0.67) | 3.52 (0.61) | 3.64 (0.55) | 3.74 (0.50) |
| PPIs | 2.53 (0.67) | 2.78 (0.72) | 2.99 (0.75) | 3.09 (0.72) | 3.25 (0.70) |
| ACE inhibitors | 3.18 (0.72) | 3.29 (0.66) | 3.40 (0.65) | 3.47 (0.61) | 3.55 (0.61) |
| Angiotensin receptor blockers | 2.52 (0.72) | 2.66 (0.70) | 2.70 (0.69) | 2.78 (0.73) | 2.97 (0.71) |
| Calcium channel blockers | 2.54 (0.87) | 2.87 (0.85) | 3.06 (0.85) | 3.13 (0.84) | 3.25 (0.84) |
| Beta blockers | 3.28 (0.66) | 3.37 (0.64) | 3.45 (0.66) | 3.44 (0.63) | 3.49 (0.69) |
| SSRIs | 3.43 (0.66) | 3.71 (0.66) | 3.86 (0.67) | 3.94 (0.68) | 4.20 (0.66) |
| Penicillins | 2.68 (0.51) | 2.87 (0.39) | 2.93 (0.34) | 2.97 (0.29) | 3.01 (0.29) |
| Opioids | 3.76 (1.53) | 4.70 (1.28) | 5.17 (1.18) | 5.43 (1.10) | 5.75 (1.04) |

**Figure 2.**
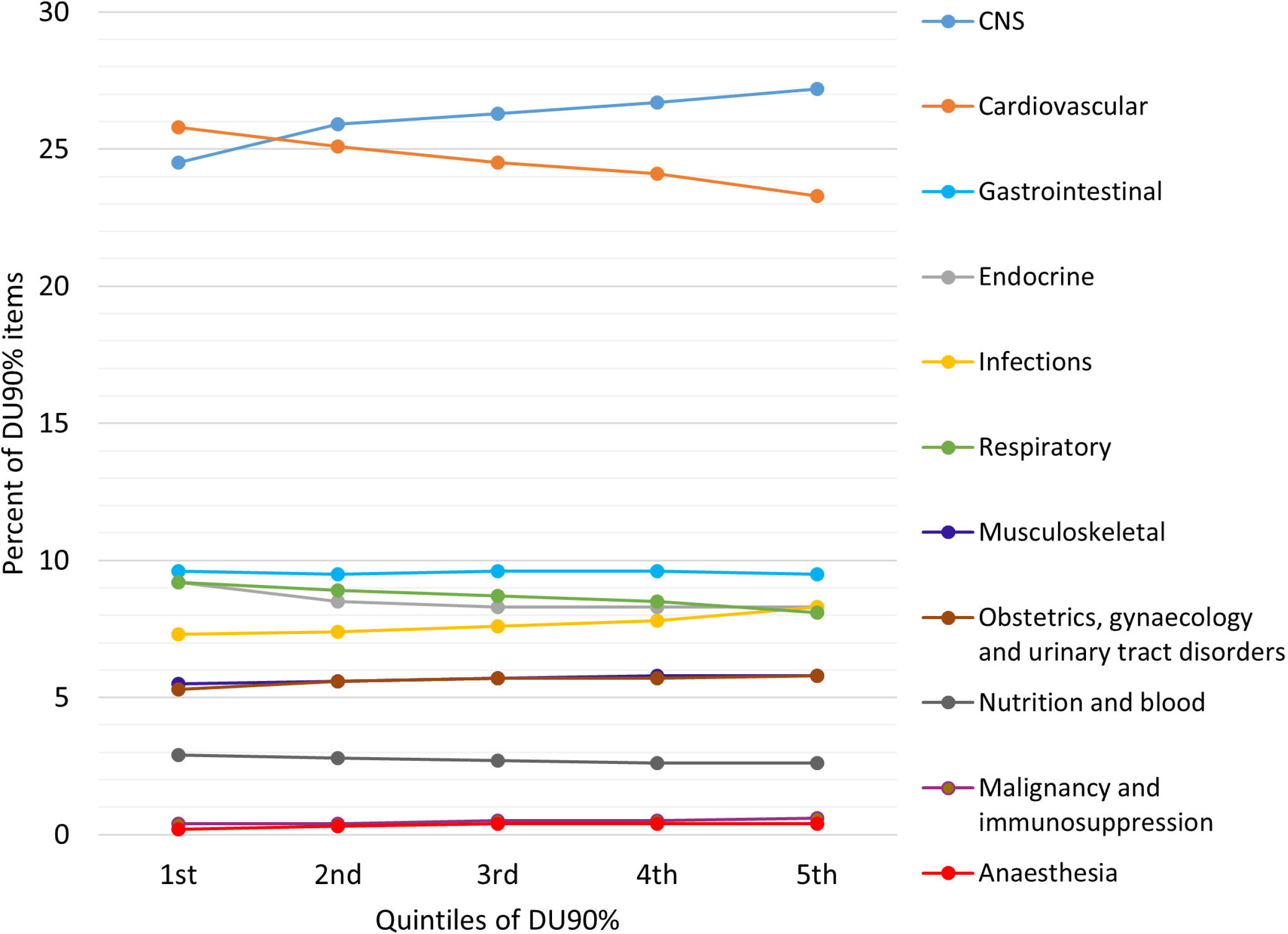
Percent of DU90% prescriptions by BNF chapter (therapeutic domain) across quintiles of DU90%.

## Discussion

Our study found that typically 130 unique medications account for 90% of the prescribing volume of GP practices in England, and variation, which was primarily between practices, was relatively low. The mean DU90% of 130 per GP practice in this study is similar to previous studies which reported means ranging from 128 to 165 drugs in practices’ DU90%, and variation similarly in the range of two fold.[3, 16, 20]

We found that greater numbers of patients and a greater proportion of patients aged 45 years and above were associated with a higher DU90%. Controlling for the number of prescriptions, a large patient population would more likely require a greater variety of medications within the DU90%. In the same way, the prevalence of multimorbidity increases with age,[21–23] and so for older people it may be more likely that they require multiple medications or thatfirst-line treatments may be unsuitable for them due to interactions with their other drugs or conditions.[24] Hence a higher proportion of older patients may also contribute to a higher DU90%. A higher proportion of female patients was also associated with a higher DU90% although to a lesser degree, again explained perhaps by research showing women are more likely to have multiple conditions and take more medicines.[22, 23] There was also a relationship between higher DU90% and high deprivation. Deprivation is associated with poorer health outcomes and greater multimorbidity,[21] and potentially less time available for GPs to optimise or rationalise prescribing due the increased health needs of the population and resulting demand.[25]

Taking practice workforce characteristics into consideration, more full time equivalent GPs and the presence of a GP registrar were both associated with a reduction in the DU90%. More GPs working in a practice and their individual prescribing habits are likely to contribute to variation in prescribing and a higher DU90%. Presence of a GP registrar indicates practices which may be more actively engaged in education and practice development. Our findings are somewhat consistent with recent research which found GP training practices had higher patient satisfaction across several domains,[26, 27] had higher QOF scores,[27] higher scores for both level and quality of services,[28] prescribed fewer antibiotics overall and broad-spectrum antibiotics.[29]

Future research could consider applying the DU90% in specific therapeutic domains. Previous domain-specific research in primary care has focussed on priority or high-risk drug classes, particularly antibiotics, where variation in the overall number of agents, broad-spectrum agents, and non-first line agents in the DU90% has been examined.[30–34] Variation in DU90% profiles of non-steroidal anti-inflammatory drugs,[35–37] and central nervous system agents have also been examined.[38, 39]

Audit and feedback is an important intervention for quality improvement.[40] Implementing the DU90% within practice management software systems or OpenPrescribing.net, for example, could provide prescribers with an opportunity to benchmark themselves against other GP practices.[41] Specifically, feedback on a practice’s DU90% within specific therapeutic domains and the agents that predominate could be used in conjunction with local formularies to identify areas where prescribing could be optimised. Previous studies have found positive attitudes among prescriber to feedback incorporating their DU90%.[16, 42, 43]

There is potential that information on medications used commonly by many practices could be considered in formulary development or updates. Previous research demonstrates that practices can vary substantially in their response to changes in prescribing recommendations.[44] Therefore, the current predominant practice may be a consideration where there is little clinical or cost-effectiveness difference between alternatives drugs. Information on predominant medications can also be useful in education. The development of a ‘starter’ formulary for trainee prescribers has been an important step, as during training, students may be exposed to varied prescribing practices.[8, 10, 11] However, even more experienced prescribers, depending on their pharmacology training,[8] could also benefit from rationalising their personal formulary to enhance quality of prescribing and potentially reduce errors. In total, approximately 600 medicines featured in the DU90% of at least one practice, representing about half of medications included. Although GPs within a single practice may be using a core group of drugs themselves, co-ordinating prescribing among their colleagues in the same practice may be beneficial where GPs may be renewing prescriptions initiated by other prescribers in the practice.[46] This is supported by evidence that the number of prescribers is an important factor in the risk of inappropriate drug combinations.[47]

While clinicians may be encouraged to focus on prescribing a smaller range of medicines, measures to reduce the variety of drugs prescribed have been applied via restricted formularies or reimbursement lists.[9] The ‘Wise List’ is a formulary of essential medicines in the Stockholm region of Sweden, and is regarded as trustworthy and useful by primary care doctors.[48] It can be regarded as effect, as adherence to its recommendations in primary care has increased from 80% in 2000 to 90% in 2015, with practice variation decreasing from 32% to 13%.[12] The ability to restrict the number of medications on the market at the level of individual countries is limited in Europe by membership of the European Union and measures such as the European Medicines Agency’s centralised approval process. For example, Sweden experienced a substantial growth in the number of pharmaceutical products on its market after joining the European Union in 1995 and adopting EU regulations.[3]

A limitation of our study is that data was available only at the GP practice level, rather than at the level of individual prescribers. Hence a high DU90% may be optimal if different prescribers in a practice have expertise that covers a wider breadth of medicines, and this is reflected in the finding of a relationship between more GPs and higher DU90%. However, there was still a relationship between higher DU90% and low value prescribing and medication costs. While previous studies have assessed the proportion of guideline recommended medications with practices’ DU90%, we were limited by the lack of national formulary. Although local formularies do exist, these are not all publicly available, or in a form while allows the information in them to be easily reused. Publication of local formularies in machine readable, interoperable formats, including coded medication details (such as BNF, ATC or SNOMED), would facilitate further drug utilisation research. A further limitation is that we lacked detailed information about the registered patient populations for each practice, and thus full adjustment for differences in case mix between practices was not possible. However, we did account for the age and sex profile of practices’ patients. Other factors such as exposure to pharmaceutical representatives,[49] or the level of prescriptions initiated in specialist care, could also explain some variation in the DU90% and prescribing costs.

This study is the first to our knowledge to examine trends in the DU90% over time, and to apply this measure in an English setting, and provides evidence that GP practices with higher DU90% tended to have higher prescribing costs and more prescribing of low value care (i.e. low effectiveness, poor cost-effectiveness, or being low priority). The DU90% could be implemented into GP practice prescribing analysis in the future and used for audit and feedback purposes to help optimise prescribing practice, through OpenPrescribing for example.[41] This could be tailored using details of local formularies to also include a measure of adherence to recommendations within the DU90% segment. Such tools which may help doctors optimise their prescribing are important to maximise medication benefits and reduce harms. This is a clear priority globally, reflected in the World Health Organisation’s current Third Global Patient Safety Challenge, ‘Medication without Harm’.[50]

## Data Availability

The data that support the findings of this study are openly available in Zenodo at https://doi.org/10.5281/zenodo.3894539.

## Author contributions

FM conceived the study, and CC, MM, TF and FM designed the study. CC collated the data and CC and FM conducted the analysis. All authors interpreted the findings. CC and FM drafted the manuscript and all authors critically revised the manuscript.

## Appendices

**eTable 1 Details of non-oral products and corresponding BNF codes excluded before DU90% calculation**

**eTable 2 List of 17 medications or classes considered as low priority items**

**eTable 3: Summary of practices’ DU90% and measures of variation**

**eTable 4: Frequency of medications contained in any practice’s DU90% segment in 2013**

**eTable 5: Frequency of medications contained in any practice’s DU90% segment in 2017**

**eTable 6: Frequency of medications contained in the DU90% segment for practices in the lowest quintile of DU90% in 2017**

**eTable 7: Frequency of medications contained in the DU90% segment for practices in the highest quintile of DU90% in 2017**

**eFigure 1 Distribution of practices across quintiles of DU90% by number of different drugs in each class included in their DU90%**

